# Enhancing Prediabetes Diagnosis from Continuous Glucose Monitoring Data via Iterative Label Cleaning and Deep Learning of Bridge2AI AI-READI Data

**DOI:** 10.64898/2026.03.04.26347604

**Authors:** Nikhil Jivraj Arethiya, Lori Krammer, John David, Vishal Bakshi, Atin BasuChoudhary, Urnisha Bhuiyan, Sabyasachi Sen, Raja Mazumder, Patrick McNeely

**Author notes:** Co-first authors (Nikhil Jivraj Arethiya, Lori Krammer).

## Abstract

As of early 2026, over 115 million US adults (more than 1 in 3) have prediabetes, a condition with an annual conversion rate of 5%–10% to type 2 diabetes. Total diabetes (diagnosed and undiagnosed) affects approximately 40.1 million Americans, or 12% of the population, with roughly 1.5 million new cases diagnosed annually. Continuous Glucose Monitoring (CGM) provides real-time, 24/7 insights into glycemic variability, detecting dangerous highs, lows, and trends that HbA1c (a 3-month average) misses. It enables, for instance, identification of nocturnal hypoglycemia or postprandial spikes, enhancing personalized, actionable treatment decisions and improving safety. The Artificial Intelligence Ready and Exploratory Atlas for Diabetes Insights (AI-READI) dataset was produced by the National Institutes of Health (NIH) Common Fund Data Ecosystem (CFDE) Bridge2AI program. This dataset offers a rich resource for diabetes research, providing comprehensive biosensor data from over 1,067 participants. However, like many medical datasets, AI-READI contains label inaccuracies due to self-reported health surveys and static HbA1c indicators, which can undermine model effectiveness. We developed a strong classification framework using Convolutional-Bidirectional Long Short-Term Memory (Conv+BiLSTM) to analyze and accurately classify glycemic health states from continuous glucose monitoring time-series data. Our aim was to establish and correct any misclassified labels through hybrid unsupervised-supervised learning methods and validated our results with expert-in-the-loop clinical review. We analyzed 784 participants from the AI-READI dataset, which represented four health states: healthy, prediabetes lifestyle controlled, oral medication, and insulin-dependent. Based on recommendations from the literature and our own expertise, we sought to compare the self-provided “healthy” group labels with a cluster-agnostic, CGM-defined healthy (CGM-H) reference derived from the CGM metrics using K-means clustering (K=6) on standardized CGM summary features to identify CGM-H participants and then applied XGBoost-based iterative label refinement. We identified a misclassification rate of 56.9% (161/283) in the initially labeled “healthy” group. After eight iterations of XGBoost refinement with dual-criterion relabeling (≥80% probability + unanimous out-of-fold voting), the cleaned dataset increased CGM-H participants from 122 to 195 for binary classification. Next, we developed a Conv+BiLSTM model combining Convolutional layers (32, 64 filters) for local temporal feature extraction with Bidirectional LSTM layers (64, 32 units) for sequence modeling, using time-series engineered features including rolling statistics, glucose derivatives, and circadian rhythm encoding. Class imbalance was addressed with per-class weighting, and 5-fold stratified cross-validation estimated generalization performance, computing a global decision threshold (0.374) by maximizing Youden’s J statistic on concatenated out-of-fold predictions. Additionally, we analyzed heart rate, activity level, and stress and sleep data and validated it against CGM data. The Conv+BiLSTM model achieved ROC-AUC ≈ 0.932 on the held-out test set and 0.907 ± 0.026 in cross-validation, with well-calibrated predictions (Expected Calibration Error = 0.075, temperature scaling T = 1.00). A 3-tier confidence-based decision system achieved 82% detection rate with only 6% OGTT burden, enabling actionable clinical recommendations. This hybrid approach addressed label noise while achieving high discrimination. This framework demonstrates potential for real-time glycemic state monitoring and early intervention in diabetes progression.

## INTRODUCTION

### Background and Clinical Motivation

Diabetes mellitus has grown to become a global health crisis, affecting over 537 million individuals globally^1^. As of early 2026, in the United States alone over 115 million US adults prediabetes, a condition with an annual conversion rate of 5%–10% to type 2 diabetes. Total diabetes affects approximately 40.1 million Americans - nearly 12% of the population - with roughly 1.5 million new cases diagnosed annually.^2^ This condition progresses along a continuum from normal glucose homeostasis to prediabetes and, in some individuals, to full diabetes. Classic estimate suggests that approximately 5–10% of individuals with prediabetes are estimated to progress to type 2 diabetes annually, with a similar proportion reverting to normoglycemia. In contrast, more contemporary pooled cohort studies showed that 12.5% progressed to type 2 diabetes within 10 years, and 36.1% reverted to normal glycaemia, with transition risk influenced by baseline fasting glucose, age, sex, and ethnicity^3, 4^. This gradual yet heterogeneous process transitions from metabolic dysregulation to chronic disease, reflecting a process of extremely complex physiological changes that are not well captured by current diagnostic markers^5^. Clinically, physicians continue to depend primarily on static biomarkers such as Hemoglobin A1 (HbA1c) and fasting plasma glucose level, and all they reveal is a snapshot without reflecting the dynamic trends of glucose and the metabolic variations that occur in the daily life of an individual^6, 7^.

The Centers for Disease Control and Prevention (CDC) has reported that over 115 million adults (about 40% of the adult population in the United States) have prediabetes^2^. Only 19% of these individuals have been told by a healthcare professional that they have prediabetes, leaving more than 80% of them unaware of their condition^8^. This combination of high prevalence and low awareness highlights a wide gap in prevention, screening, and early intervention efforts. The introduction of continuous glucose monitoring (CGM) technology has completely changed how we can approach diabetes management. With these sophisticated devices, we receive real-time glucose measurements every 5 minutes, which provides us with data for analysis that is exceptionally rich^9^. This temporal detail allows us to detect glucose variability patterns, calculate time-in-range metrics, cooling periods, and even identify glycemic excursions, factors that we now know strongly correlate with long-term complications of overall patient health^7, 10, 11^. The challenge is that converting all this CGM data into something clinically meaningful calls for advanced analytical techniques that can handle the noise and complexity that come with time-series biological data, while keeping it interpretable enough for medical professionals to make clinical judgments^12^.

We found a significant gap between the amount of data CGM devices can provide and our ability to turn it into useful therapeutic insights. Although CGM technology has advanced rapidly, the statistical methods for handling and interpreting this data have not kept pace, leaving doctors with too much data and not enough ways to use it to improve patient care^7, 12–14^.

### Continuous Glucose Monitoring (CGM) data

Prior studies in CGM data analysis and classification have concentrated on either glucose prediction or pattern recognition using standard ML techniques^15, 16^. Time-series modeling approaches for glucose prediction have been promising, yet most studies presume accurate ground truth labels and do not consider the inherent noise within medical datasets^17^. The problems of label noise in medical datasets are a pressing but underappreciated concern in diabetes research^18^. In summary, predictive pipelines for CGM frequently assume labels are fixed when they are uncertain in the real world, which can bias model selection, calibration, and subgroup performance; this is the gap for label-aware evaluation and curation in advance of and/or parallel with modeling.

One major strand of research has focused on CGM-derived variability metrics and their clinical relevance^19, 20^. Time-in-range (TIR) and its corresponding measures (time-below-range (TBR), time-above-range (TAR)) are now the defined clinical endpoint for understanding CGM in both clinical and research scenarios^21^. The Advanced Technologies & Treatments for Diabetes (ATTD)/American Diabetes Association (ADA) international consensus statement recommends that clinicians report TIR as a standardized target and contextual measure, making TIR actionable as a proxy measuring overall glycemic control along with HbA1c to direct treatment decisions. When this label was first published in 2017 and then included within a revised set of statements published in 2019, it highlighted the importance of out-of-range measures and harmonizing reports of CGM across populations and trials. Together, these statements fundamentally shifted CGM from high-frequency, raw data to standardized decision support summaries, facilitating both comparison and any downstream analytics^6, 7^.

### Glycemic Variability

In addition to TIR/TBR/TAR, glycemic variability (GV) describes short- and medium-term glucose variation and its contribution to complications risk and patient-reported outcomes^22, 23^. Classic GV metrics include the coefficient of variation of glucose (%CV), Mean Amplitude of Glycemic Excursions (MAGE), Continuous Overlapping Net Glycemic Action (CONGA), and the J-index^24–26^. The %CV measure is easy to compute, and performance analysis shows it is robust; a widely used threshold near 36% separates relatively stable from unstable glycemia^20^. MAGE summarizes the magnitude of clinically meaningful excursions, CONGA captures the intraday dispersion of glucose in hour-specific time-lags (e.g., 1-2 hrs.), and the J-index combines the mean level with dispersion into a single control score, metrics that complement TIR/%CV rather than replace them.

In our context, this combination (TIR/TBR/TAR, %CV, MAGE, CONGA, J-index) is appealing because (i) it aligns with international reporting standards (TIR/%CV); (ii) it encompasses excursion size (MAGE); (iii) it provides a measure of temporal instability at clinically meaningful horizons (CONGA 1-2 hours); and (iv) it encapsulates overall control (J-index), which directly serves two of our downstream purposes: to triage CGM healthy (CGM-H) in contrast to variability (high TIR, low variability) and also identify labeling that is inconsistent with physiological evidence.

At the modeling level, we therefore engineer time-series features that retain physiological structure while mitigating sensor noise: short-window rolling statistics (mean/standard deviation (SD) within a 1-h epoch), first and second derivatives (the rate of change and acceleration), circadian encodings (sin_hour, cos_hour), and contextual flags (is_night, is_meal_time, is_sleep). These are all consistent with common reporting practices and normative CGM profiles aggregated from healthy cohorts, providing interpretable predictors for clustering, relabeling, and Convolutional-Bidirectional Long Short-Term Memory (Conv+BiLSTM) modeling^7, 27^.

Understanding postprandial kinetics is an essential component of assessing glucose “cooling periods” after a spike. In metabolically healthy people, glucose is usually at its peak within 30–60 minutes after eating and returns to baseline within 2-3 hours^10, 28^.

Dysglycemia is an umbrella term for any abnormality in blood sugar regulation, encompassing both hyperglycemia (high blood sugar/diabetes) and hypoglycemia (low blood sugar). It indicates unstable blood sugar, often stemming from insulin resistance or pancreatic issues, resulting in fatigue, shakiness, confusion, and increased cardiovascular risk. Management involves diet changes, regular exercise, and medication^29^.

In prediabetes and dysglycemic states, glucose levels peak higher, return to baseline more delayed, and remain in hyperglycemia phases for longer beyond the baseline, often returning to baseline after three hours. These patterns provide a physiological anchor for interpreting recovery metrics in CGM. And when paired with the excursion segmentation, they help distinguish between an isolated spike and prolonged hyperglycemia. Our analysis used these biomarker benchmarks as anchors to interpret whether the prolonged degree of recovery time in our cohort truly indicated dysglycemia or an artifact from the algorithm.

### Dataset Challenges and Research Gaps

The AI-READI (Artificial Intelligence Ready and Exploratory Atlas for Diabetes Insights) dataset, supported by the National Institutes of Health (NIH) Common Fund Data Ecosystem (CFDE) Bridge2AI funding, is a flagship dataset for diabetes research that includes over 1,000 participants with extensive biosensor data^30^. The dataset was carefully constructed to fast-track AI-driven discoveries for diabetes care by using systematic data collection protocols. Nevertheless, the participant health status labels were primarily derived from self-reported questionnaires and single-timepoint (static) HbA1c measurements, resulting in noticeable label noise that degrades supervised learning performance and, by extension, potentially affects clinical safety.

Our initial analysis revealed several concerns: (i) inconsistencies in the “healthy” group, with some participants displaying glucose patterns more consistent with prediabetes, (ii) the presence of non-numeric sensor outputs such as “LOW” and “HIGH,” and (iii) uneven distribution of participants across health states. These issues highlight the challenge of working with noisy, heterogeneous medical datasets. In medical datasets, label noise is prevalent because the determination of ground truth often relies on subjective reporting, surrogate markers (e.g., HbA1c alone), or outdated evaluation methods^5, 7, 31^. This has major consequences not only for model performance but also for clinical safety, as misclassified persons may be given inappropriate recommendations, or miss opportunities for early intervention.

In addition to dataset-related challenges, CGM-based classification strategies are further confronted with three persistent, field-wide challenges: (1) label quality and expert-in-the-loop relabeling remain under-developed in practice, even though systematic evidence shows that noisy labels impair medical ML performance, and targeted “active” cleaning substantially improves it^31,32^; (2) sequence-aware, clinically interpretable feature engineering for CGM (derivatives, circadian encodings, recovery dynamics) lacks standardized outside of the consensus summary metrics (e.g. TIR and % CV)^7, 20^; and (3) there is no consensus on minimum CGM data requirements for robust modeling, despite clinical guidance favoring ∼14 consecutive days with ≥70% wear and recent studies questioning whether longer windows are needed for some endpoints^33, 34^. These challenges motivate our approach and frame the scope of the study.

In response, we deliver four integrated contributions: (1) a label-aware, expert-in-the-loop hybrid unsupervised-supervised approach for label refinement that can be applied broadly to medical datasets while avoiding information leakage, (2) a comprehensive feature engineering framework specifically designed for CGM time-series analysis, (3) determination of minimum data requirements for reliable classification, and (4) an open-source methodology for improving medical dataset quality that promotes reproducible research. We prioritized participant-level splits and training-only standardization to minimize leakage and optimistic bias in all downstream analyses. Our pipeline combines unsupervised CGM-phenotype audits to identify implausible “healthy” labels with an iterative feedback loop involving XGBoost relabeling and expert assessment^31, 35^. This hybrid approach ensures that labels are clinically coherent before training the downstream LSTM model on carefully selected, physiologically relevant features^36^. This methodology builds upon our existing work^37–42^.

## MATERIALS AND METHODS

### Dataset Description and Preprocessing

The AI-READI included 1,067 participants, of whom 1,049 had continuous glucose monitoring (CGM) data recorded using Dexcom G6 sensors at 5-minute intervals over 2-13 days. Each participant’s data was stored in an independent JSON file having structured glucose readings with timestamps, device information, and metadata. The dataset consisted of four categories of health states: healthy (n=372), prediabetes-lifestyle-controlled (n=242), oral medication-dependent (n=323), and insulin-dependent (n=130). Our initial investigation revealed significant problems with the quality of data that required methodical preparation. Several (n=240) participants contained “LOW” and “HIGH” string values rather than numerical glucose values, reflecting sensor limitations or errors in measurements. These non-numerical values appeared randomly during monitoring sessions, not just at start/end points. These participants were removed from our final analysis. For reproducibility, we flattened each participant’s JSON to a row-wise table, harmonized labels to a consistent four-class schema, and exported participant chunks into batched CSVs prior to feature engineering.

To provide sufficient temporal coverage for reliable pattern detection, we set a minimum threshold of 2,138 CGM record samples (≈7 days at 5-minute cadence) based on the percentile analysis of the dataset. This cutoff included 784 participants (≈97th percentile) and preserved enough data density for stable time-series modeling. We prioritized completeness of data over sample size to guarantee that our models would have trustworthy training data. The parsing step involved the extraction of glucose readings, timestamps, and participant metadata during the traversal of the nested JSON structures of the raw files. Temporal alignment was carried out to normalize all sequences to a fixed length of 2,138 records to enable meaningful analyses across all participants. We employed masking strategies in our downstream modeling strategies to naturally handle variable-length sequences while preserving the original temporal patterns. Any individual traces that were greater than the fixed length window were retained only for their earliest window, and any shorter traces were excluded upstream to prevent imputation artifacts from rejections, consistent with the related codebase filters.

### Feature Engineering Framework

#### Clinical Glycemic Metrics

We calculated conventional clinical measurements such as TIR (70-140 mg/dL for non-diabetic physiology), TAR (>140 mg/dL), TBR (<70 mg/dL), mean glucose levels, SD, and coefficient of variation. These measurements provide a valuable glucose control review following current diabetes care approaches^43^. These per-participant metrics were later used both for phenotype clustering and for the supervised relabel loop.

#### Advanced Glycemic Variability Metrics

Other metrics of variability included MAGE for quantifying the magnitude of glucose excursions, CONGA for assessing temporal stability, J-Index for quantitation of overall variability, and Lability Index^44^ for measuring rapid fluctuations. MAGE and CONGA were computed from the 2,138-point sequences, with CONGA evaluated at one- to two-hour lags to align with intraday dynamics.

#### Time-Series Specific Features

We extracted temporal features for Conv+BiLSTM modeling^45^, such as glucose change rate (first derivative), glucose acceleration (second derivative), rolling statistics (1-hour rolling mean and SD), temporal encoding (*hour*, *day fraction*, *sin_hour*, *cos_hour*), and behavioral indicators (*is_night* (10 PM-6 AM), *is_meal_time* (3-hour windows around typical meal times like 7-9 AM, 12-2 PM, and 6-8 PM), *is_sleep* (11 PM-7 AM)). These features represent circadian rhythms and glucose dynamics, which are required for accurate classification^46^. In practical terms, the 1-hour windows applied in the analysis were determined as a 12-sample (5-min cadence) rolling mean/SD, *glucose_diff* identified the first-differences in glucose, and cycles of time coding which are sin/cos of hour were used to convey circularity. For the final Conv+BiLSTM model, the selected input features were: blood_glucose_value, glucose_rollmean_1h, glucose_rollstd_1h, glucose_diff, glucose_accel, sin_hour, cos_hour, is_meal_time, and is_night (9 features total); other engineered variables were retained for analysis but not fed to the network. Continuous features were standardized using train-only statistics; binary flags were left unscaled.

### Label Quality Assessment and Clinical Validation

#### Clinical Consultation

Scientific evidence, augmented by consultation with clinical endocrinologists, indicates that CGM-H individuals ought to spend more than 90% of their time in the range (70-140 mg/dL) with minimum glycemic variability^7, 43^. Visual examination of glucose patterns from participants classified as “healthy” revealed patterns that questioned normal glucose equilibrium, requiring thorough label verification. Accordingly, we formalized a two-phase audit to (i) discover CGM phenotypes in the nominally healthy group and (ii) iteratively reconcile labels under expert review. All expert review in this work was done in collaboration with one of the authors, Sabyasachi Sen, MD, currently Chief of the Division of Endocrinology at the Veterans Affairs Medical Center in Washington D.C.

#### Phase 1: Unsupervised Clustering

K-means clustering was applied using the scikit-learn implementation^47^ to the originally labeled healthy group (n=283 after data cleaning) on standardized summary features: *mean_blood_glucose*, *std_dev*, *cv_percent*, *tir_percent*, *tbr_percent*, *mage*, *lability_index*, and *conga_2h*. Features were z-normalized to ensure equal contribution. The optimal cluster number was determined through systematic evaluation of K=2 to K=10 using three complementary metrics: (1) Elbow method using KneeLocator (Fig. 1A), (2) Davies-Bouldin Index (measures cluster compactness; lower is better), and (3) Silhouette analysis^48^ score (measures cluster separation quality). K=6 was selected based on convergence of elbow method and Davies-Bouldin Index minimization, balancing statistical validity with clinical interpretability. We illustrated the cluster structure via Principal Component Analysis (PCA) projection of the standardized features (PC1=59.2% variance; PC2=22.1%), which depicts the CGM-H cluster as well-defined in nature and separated from the higher variability groups (Fig. 1B). A summary of these features, their clinical rationale, and their downstream use is shown in Table 1. To characterize “healthy” in a cluster-agnostic way, we chose the cluster with the highest TIR, lowest mean glucose, and lowest %CV. The cluster ID is data-dependent and recorded in the audit log.

**Figure 1.**
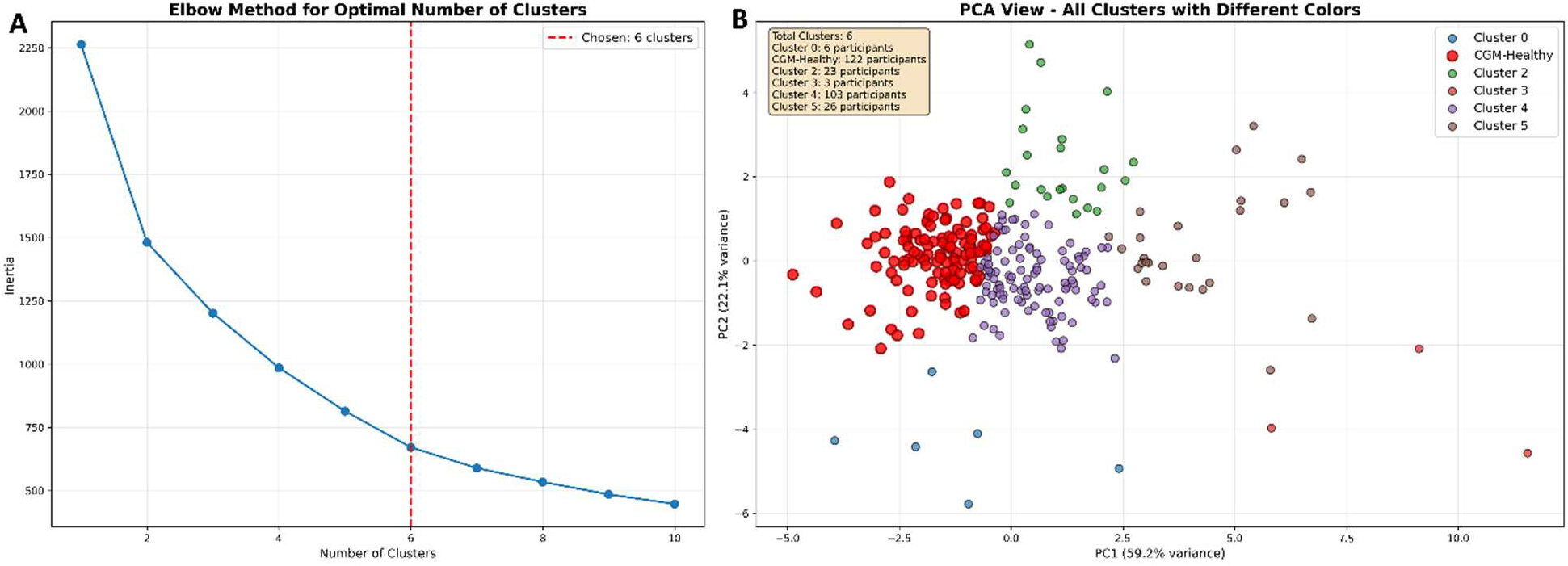
Cluster selection and structure. (A) Elbow plot of K-means inertia identifying K=5 at the inflection point. (B) PCA projection (PC1=70.1% variance; PC2=19.5%, the standardized CGM summary features colored by cluster); the CGM-Healthy group (red) has a generally compact dispersion and shows clear separation from the other clusters with greater variability.

**Table 1.** Summary of features used in clustering, their rationale, and downstream modeling use.

| Feature | Clinical Rationale |
| --- | --- |
| *mean_blood_glucose* | Represents overall glycemic exposure; elevated mean values are indicative of dysglycemia and increased long-term risk |
| *std_dev* | Quantifies dispersion of glucose values, reflecting overall variability in glycemic control |
| *cv_percent* (%CV) | Normalized variability metric; values >36% are associated with unstable glycemia in consensus reports |
| *tir_percent* | Internationally recommended marker of glycemic control; >90% time in range is typical in metabolically healthy individuals |
| *tbr_percent* | Captures hypoglycemia burden, a clinically critical safety endpoint |
| *mage* | Mean Amplitude of Glycemic Excursions; summarizes the size of clinically significant postprandial spikes |
| *lability_index* | Reflects frequency and magnitude of rapid intra-day glucose swings |
| *conga_2h* | Continuous Overlapping Net Glycemic Action; measures stability across 2-hour lags, capturing intraday dispersion |

We examined glucose profiles for each individual in every cluster to validate cluster assignments against clinically meaningful CGM patterns. The cluster meeting the predefined criteria (highest TIR; lowest mean and %CV) was designated as CGM-Healthy and consistently showed normal glucose patterns, with high time in range (over 90%) and low variability. In contrast, the other clusters displayed prediabetes features, including high average glucose levels, high variability, and low TIR. We retained cluster membership and the review decision in an audit log to support downstream supervised refinement.

#### Phase 2: Supervised Iterative Refinement

XGBoost models were trained using the recently cleaned labels to find additional misclassifications across all health states^35^. We followed an iterative procedure: first, we performed a participant-level train/test split (70/30, stratified) before any resampling, then we created a frozen test set of 30% that remained untouched throughout all iterations to prevent data leakage and ensure unbiased evaluation. Synthetic Neighbor Oversampling Technique (SMOTE)^49^ was applied to the training fold only, and the test fold was left untouched. At each round, we trained XGBoost on the current labels using 15 stratified out-of-fold (OOF) splits with 3 independent predictions per participant, identified high-confidence cases predicted as CGM-defined healthy (CGM-H) with ≥80% mean probability and unanimous voting (3/3 OOF predictions) but not labeled so, and assessed these candidates against clinically informed criteria established in consultation with our expert. Expert accept/reject decisions were written back to a shared label file. We repeated this process in iteration until it converged with no participant left to be relabeled. To limit leakage, all resampling was restricted to train only, candidate relabels were proposed at the participant level, and each round recorded (i) the number flagged, (ii) expert accept/reject decisions, and (iii) the net class flow. The iterative process continued until either (1) fewer than 10 new relabeling candidates emerged in a single iteration, or (2) validation AUC changed by less than 0.5% between consecutive rounds. Candidates were flagged using a dual criterion of ≥80% mean probability and unanimous out-of-fold voting (3/3 OOF predictions). The frozen test set of 30% remained unchanged throughout all iterations to prevent data leakage.

### Cooling-Period Analysis

We measured post-excursion recovery (“cooling”) with a baseline and living range determined separately for each participant. First, we estimated baseline and living range from the glucose distribution using four complementary views: (i) percentiles 10th–90th (central 80%), (ii) percentiles 15th–85th (central 70%), (iii) a histogram-mode approach (100 bins; modal bin ± neighboring bins with ≥20% of peak density), and (iv) a two-component Gaussian mixture where the heavier component defined the living mode with bounds at mean ±2.5 SD^50^. Candidate baselines/ranges outside physiologic limits (baseline 60–250 mg/dL; width 10–150 mg/dL) were discarded, and a consensus baseline and upper bound were taken as the median across valid methods.

Spikes were detected with threshold height ≥ living-range upper, minimum inter-peak distance of 3 samples (15 minutes), and prominence ≥ 5 mg/dL. The primary recovery endpoint was the sustained return time: from the peak index to the first time the glucose fell at or below the living-range upper and remained there for ≥3 consecutive samples (≥15 minutes). Requiring ≥3 consecutive readings helped to avoid false drops caused by brief fluctuations during recovery. Episodes without a sustained return within 6 hours were treated as right-censored. We also computed an exploratory 80%-recovery time (baseline + 0.2 × peak-to-baseline amplitude), reported descriptively. Because CGM captures interstitial rather than capillary glucose, an intrinsic 5–10-minute lag is expected^51^; the sustained-return criterion mitigates spurious one-point dips during the decay.

### Conv+BiLSTM Classification Model Development

The Conv+BiLSTM (Convolutional-Bidirectional Long Short-Term Memory) model^52, 53^ was designed to address the challenge of long CGM sequences (2138 timesteps, ≈7.4 days) while maintaining gradient flow for effective training. The architecture comprises five key components:

1. Convolutional Frontend: Two Conv1D layers (32 and 64 filters, kernel size 3, ReLU activation) with MaxPooling (pool size 2) reduce the sequence length from 2138 to approximately 133 timesteps, extracting local temporal patterns (meal responses, glucose excursions) while compressing the sequence within the LSTM’s effective gradient-propagation range (≈200 timesteps).
2. Bidirectional LSTM Layers: Two BiLSTM layers (64 and 32 units per direction, totaling 128 and 64 bidirectional units) process the reduced sequence in both forward and backward directions, capturing context from both past and future timestamps^52^.
3. Dense Classification Layers: A Dense layer (32 units, ReLU activation) performs feature transformation, followed by Dropout (rate=0.3) and a sigmoid output layer for binary classification between CGM-Healthy and prediabetes states, implemented in Keras/TensorFlow^54^.
4. Regularization: L2 regularization (λ=0.001) was applied to LSTM kernel weights to reduce overfitting^55^. Gradient clipping (clipnorm=1.0) stabilized training on long sequences by preventing exploding gradients^56^. Dropout (rate=0.3) was applied after the dense layer.
5. Training Configuration: Optimization used the Adam optimizer (learning_rate=0.001) with binary cross-entropy loss^56^. Class imbalance was addressed using class weighting (computed as weight[class] = n_samples / (n_classes × n_samples_class)), assigning higher loss weights to the minority class (CGM-Healthy) without oversampling to preserve sequence integrity. Early stopping monitored validation loss (patience=10) with best-model checkpointing. ReduceLROnPlateau reduced learning rate when validation loss plateaued (factor=0.5, patience=5). Training used maximum 30 epochs and batch size=16^55, 56^. The complete architecture contains 121,729 trainable parameters.

Nine features were selected as Conv+BiLSTM inputs on the basis of their clinical relevance and temporal nature: *blood_glucose_value, glucose_rollmean_1h, glucose_rollstd_1h, glucose_diff, glucose_accel, sin_hour, cos_hour, is_meal_time, and is_night.* These features provide a reasonable tradeoff between model complexity and interpretability, while still capturing the important glucose dynamics. All continuous input features (7 features) were standardized using z-score normalization (mean=0, std=1) based on statistics calculated from the training portion only to prevent data leakage, and the binary flags (2 features) were left unscaled to ensure interpretability.

At the participant level, we divided the 498-participant binary classification dataset (prediabetes vs. CGM-Healthy) into 60% of training set, 20% of validation set, and 20% of held-out test set using stratified sampling to maintain class balance. To robustly estimate generalization performance, we implemented 5-fold stratified cross-validation on the combined training and validation sets (398 participants total), with the held-out test set remaining frozen and unseen during all training and validation procedures. In this sequence model, we did not use SMOTE or oversampling methods; instead, we addressed class imbalance using class-weighted training computed from the train labels. For each cross-validation fold, we computed the optimal threshold using Youden’s J statistic^57^ on out-of-fold validation predictions (per-fold thresholds ranged 0.10–0.44). To derive a single production-ready threshold, we re-evaluate Youden’s J on concatenated out-of-fold predictions from all five folds, yielding a global decision threshold. This global threshold was used for all subsequent held-out test set evaluations to mitigate the risk of data leakage and threshold overfitting to individual folds. Each participant’s time-series remained whole throughout training and evaluation to maintain temporal integrity.

### Post-hoc Probability Calibration via Temperature Scaling

To ensure predicted probabilities accurately reflect true class membership likelihood (essential for confidence-based clinical decision-making), we applied temperature scaling, a post-hoc calibration method that rescales model logits before applying the sigmoid function^58, 59^. Temperature scaling learns a single scalar parameter T (temperature) that adjusts probability distribution sharpness:

calibrated_probability = sigmoid(logit / T)

where T > 1 smooths probabilities toward 0.5, and T < 1 sharpens probabilities toward 0 or 1. The temperature parameter was optimized on the validation set by minimizing negative log-likelihood.

To translate model predictions into actionable clinical recommendations under uncertainty, we developed a 3-tier confidence-based decision system using dynamic thresholds centered on the global decision threshold. The system categorizes predictions into three zones, each mapped to a specific clinical action: Zone 1 (high confidence prediabetes) with predicted probability < 0.295 (global threshold − 0.08 margin), where the clinical action is immediate lifestyle intervention (diet, exercise, monitoring) without requiring confirmatory oral glucose tolerance test (OGTT)^5^; Zone 2 (uncertain) with predicted probability 0.295 to 0.595 (global threshold ± margins, width = 0.30), where OGTT is recommended for confirmatory diagnosis, capturing ambiguous cases where the model lacks sufficient confidence; and Zone 3 (high confidence CGM-Healthy) with predicted probability ≥ 0.595 (global threshold + 0.22 margin), where the clinical action is regular screening cycle (1-2 years) with no immediate intervention. Threshold margins (−0.08 for low, +0.22 for high) were selected to balance detection rate, OGTT burden, and false positive rate, and are adjustable based on local clinical workflows.

### Model Evaluation and Interpretability

The model evaluation was performed on a separate test set, using 5-fold stratified cross-validation on the training + validation set and a separate held-out test set of 20% participants, using a global decision threshold derived by maximizing Youden’s J statistic on concatenated out-of-fold predictions from all cross-validation folds. We reported ROC-AUC and PR-AUC (average precision) along with accuracy, precision, recall, F1-score, balanced accuracy, and Matthews Correlation Coefficient (MCC)^57^. Calibration was assessed using temperature scaling (post-hoc calibration fitted on validation data), a calibration curve^58^, Brier score^59^, and Expected Calibration Error (ECE)^58^, which assessed how well predicted probabilities corresponded with observed outcomes. Errors per class were summarized via confusion matrices. The splits were all at the participant level to account for leakage, and all metrics were reported both as cross-validation averages (mean ± standard deviation across 5 folds) and on the held-out test set to assess generalization performance and robustness. For interpretability, we applied sequence-aware permutation importance, in which we permuted the entire feature sequence across participants and reported ΔAUC thereafter to quantify the contribution of the respective variable^60, 61^.

For the 3-tier confidence-based decision system, we reported per-zone performance metrics including detection rate, false positive rate, and OGTT burden (percentage of cases requiring confirmatory testing). For the cooling-period analysis, we reported per-participant medians (interquartile ranges (IQR)) for completed recoveries and the proportion of right-censored episodes (no return within six hours).

We chose these metrics to ensure alignment with diagnostic machine learning best practices and account for class imbalance^62^. The ROC-AUC metric provides threshold-independent discrimination across the full operating characteristic^63, 64^, whereas PR-AUC will be more informative in the case of class imbalance as it emphasizes the precision–recall trade-off in situations with rare positive classes^65^. We add precision (positive predictive value) as a measure of false positive burden, which is important if unnecessary follow up has cost or risk, as well as the recall (sensitivity) as a measure of the risk of missed case that occurs during a screening; the F1 score provides a single summary of that trade-off. Since class sizes can be asymmetric, the Matthews correlation coefficient (MCC) and balanced accuracy (BA) metrics are more robust summaries than classification accuracy, MCC incorporates over the four cells of the confusion-matrix, and it is less sensitive to prevalence^66^ while BA averages over sensitivity and specificity. Youden’s J was selected as a clinically interpretable operating point that maximizes overall sensitivity and specificity^57^. Lastly, calibrated models are important for clinical decision support because reliable probabilities, not just hard labels, for clinical decision support are needed; Brier score reflects probabilistic accuracy and ECE measures miscalibration by bin^58, 59^.

## RESULTS

### CGM Summary Metrics

Figure 2 illustrates CGM summary metric relationships and study group separation. The correlation matrix (Fig. 2A) reveals strong associations among glucose elevation metrics (mean_blood_glucose ↔ tar_percent: r = 0.96) and variability metrics (std_dev ↔ mage: r = 0.94), with TIR negatively correlated with elevation (tir_percent ↔ mean_blood_glucose: r = −0.96). TIR distributions by study group (Fig. 2B) show clear separation: CGM-Healthy peaked at 80.6%, Prediabetes at 73.8%, oral medication at 49.8%, and insulin-dependent at 40.5%. Notably, the originally labeled “healthy” group exhibited a bimodal distribution spanning 40–95% TIR, confirming substantial label noise and justifying iterative refinement. These summary metrics informed clustering and XGBoost relabeling; the Conv+BiLSTM model subsequently learned from time-series engineered features.

**Figure 2.**
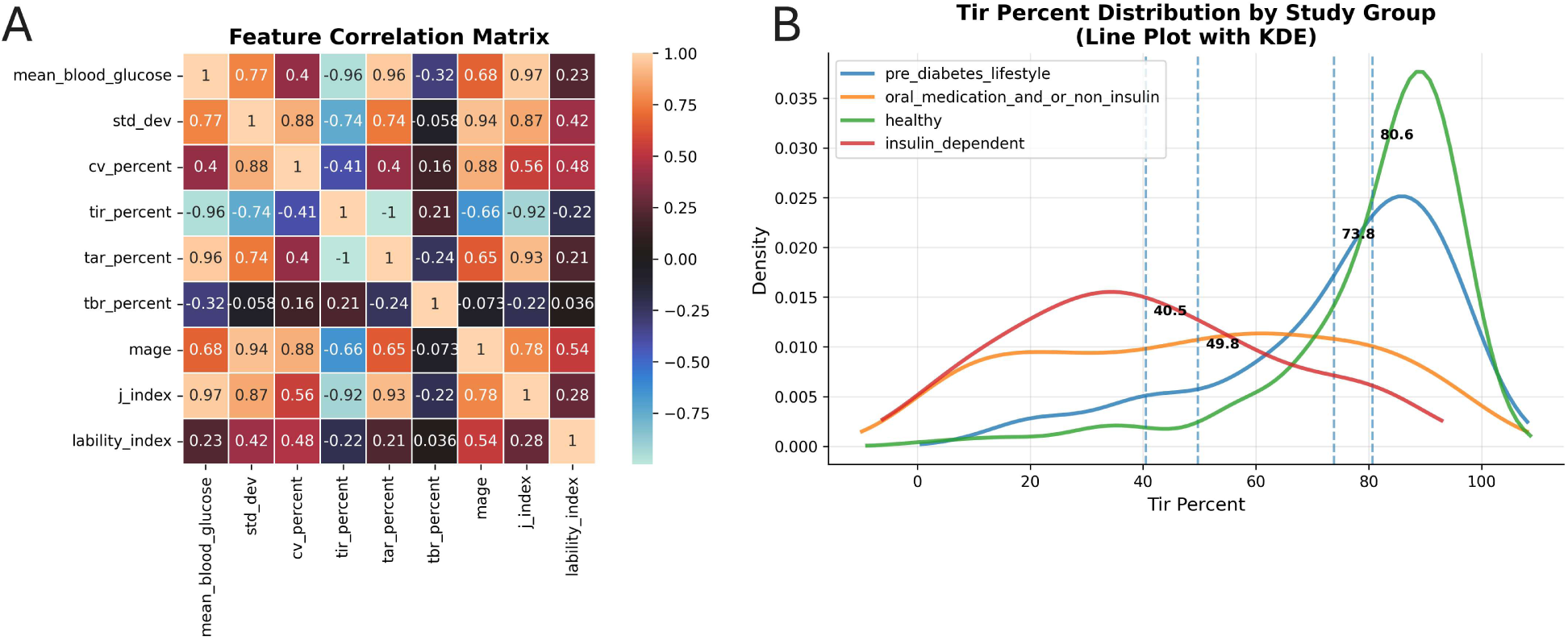
Feature correlation and distribution. (A) Heatmap matrix illustrating correlations between features. (B) Time-in-range (TIR) percent distribution for each study group.

### Label Refinement Outcomes

Results of the K-means clustering (K=6, selected via elbow method and Davies-Bouldin Index minimization) of the previously identified healthy group (n=283) indicated substantial variation in glucose patterns (Fig. 3). 122 participants (43.1%) exhibited completely good glucose control features, whereas 161 participants (56.9%) showed patterns more akin to prediabetes or early diabetes conditions. This data indicates a significant 56.9% misclassification rate in the initial healthy group. When performed, the silhouette analysis revealed that the clustering quality was robust and silhouette analysis indicated fair separation (s = 0.318) for the overall study sample and a highest per-cluster silhouette score for the CGM-H cluster (0.371), indicating superior internal compactness among all six clusters. Manual clinical validation by domain experts confirmed that the cluster selected by our predefined rule (highest TIR; lowest mean and %CV) contained physiologically healthy glucose profiles, with a TIR of 90.1% ± 6.4% (meeting the >90% criterion), mean glucose 115.2 mg/dL, coefficient of variation (CV) of 14.6%, MAGE of 32.7, and few minimal glycemic fluctuations. The CGM-healthy cluster demonstrated 16.6% higher TIR compared to other groups (90.1% vs. 73.5% ± 19.5%), confirming clinically meaningful separation.

**Figure 3.**
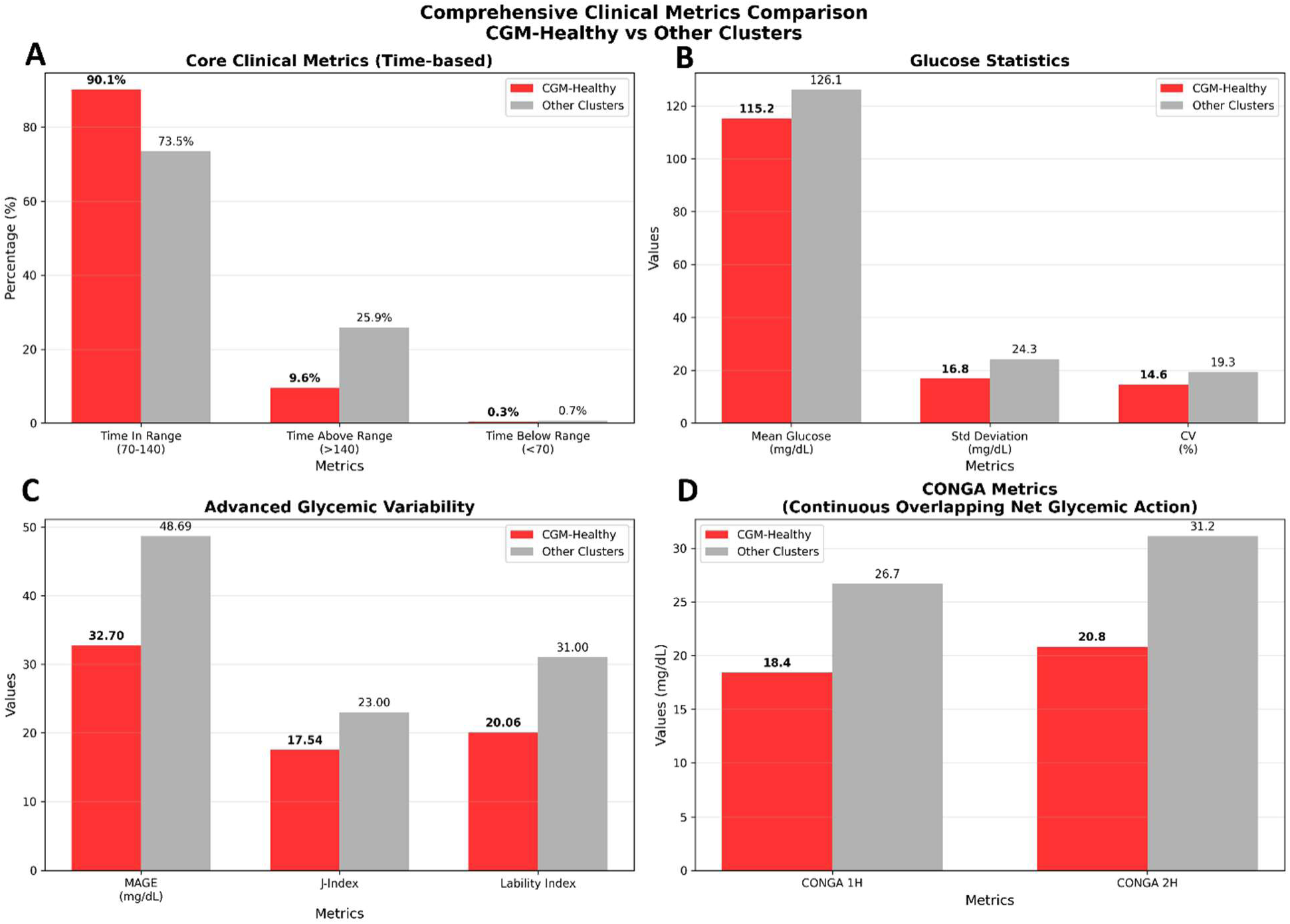
Comprehensive Clinical Metrics Comparison. (A) Core clinical metrics include time-in-range (TIR), time-above range (TAR), and time-below-range (TBR). (B) Glucose statistics, including mean, standard deviation, and coefficient of variation. (C) Advanced Glycemic Variability includes mean amplitude of Glycemic Excursions (MAGE), which indicates average size of glucose swings, with high MAGE indicating frequent or large spikes or drops, J-index, and lability index. (D) Continuous Overlapping Net Glycemic Action (CONGA) includes 1-hour and 2-hour glucose differences.

After eight iterations of relabeling refinement using XGBoost with expert review, we identified an increased number of misclassified participants across all health groups using a dual criterion of ≥80% mean probability and unanimous out-of-fold voting (3/3 OOF predictions). Convergence was achieved after iteration 8 when zero new candidates emerged. The final converged labels were validated using 5-fold cross-validation on 548 training participants (4-class problem: CGM-Healthy, prediabetes lifestyle, oral medication, insulin-dependent), achieving macro-averaged performance of ROC-AUC of 0.873 ± 0.038 (range: 0.809-0.926 across folds), Accuracy of 0.719 ± 0.043, Precision of 0.657 ± 0.068, Recall of 0.659 ± 0.061, and F1-Score of 0.653 ± 0.062. Performance was consistent across folds, with the best fold achieving ROC-AUC of 0.926 (Fold 4) and the weakest achieving ROC-AUC=0.809 (Fold 1), demonstrating robust discrimination across health states. Iterative relabeling was motivated by domain insight from our expert, who identified the plausibility of participant misclassification given the clinical heterogeneity of the cohort. Candidate relabeling were subsequently confirmed through probabilistic and voting-based analytical criteria, rather than independent clinical re-examination of each individual participants.. The iterative process increased CGM-Healthy participants from 122 (initial K=6 clustering) to 195 (final dataset for binary classification), representing a net increase of 73 participants (+59.8%). The final binary classification dataset consisted of 498 participants: 303 prediabetes and 195 CGM-Healthy, resulting in an increase in the genuine healthy cohort.

### Conv+BiLSTM Classification Performance

The Conv+BiLSTM classifier demonstrated strong performance across both cross-validation and held-out test evaluation. Stratified 5-fold Cross-validation on 398 participants achieved mean ROC-AUC of 0.907 ± 0.026, mean PR-AUC of 0.840 ± 0.040, and mean Accuracy of 0.840 ± 0.040, with consistent performance across all folds (ROC-AUC range: 0.857-0.927), confirming model stability and robustness to different train/validation splits. Per-fold optimal thresholds ranged from 0.10 to 0.44 (computed via Youden’s J on validation folds), but the global threshold strategy (0.375) reduced threshold instability while maintaining strong performance.

The model achieved excellent discrimination on the held-out test set (Fig. 4) with 20% of the data, about 100 participants were evaluated using the fixed global decision threshold of 0.375 derived from cross-validation, the model achieved an ROC-AUC of 0.932, PR-AUC of 0.848, Accuracy of 0.840, Balanced Accuracy of 0.864, and Matthews Correlation Coefficient (MCC) of 0.712. Per-class performance revealed high precision for prediabetes classification (Precision ≈ 97.9%, Recall ≈ 75.4%, F1 ≈ 85.2%), minimizing false positive burden in screening contexts, alongside high sensitivity for CGM-Healthy detection (Recall ≈ 97.4%, Precision ≈ 71.7%, F1 ≈ 82.6%), effectively capturing true healthy individuals. The model showed strong performance in both classes, effectively avoiding bias toward the majority class (prediabetes). This balance is important for fair and accurate medical diagnosis and helps boost the model’s reliability in clinical settings where both false positives (unnecessary interventions) and false negatives (missed prediabetes cases) carry clinical and economic consequences.

**Figure 4.**
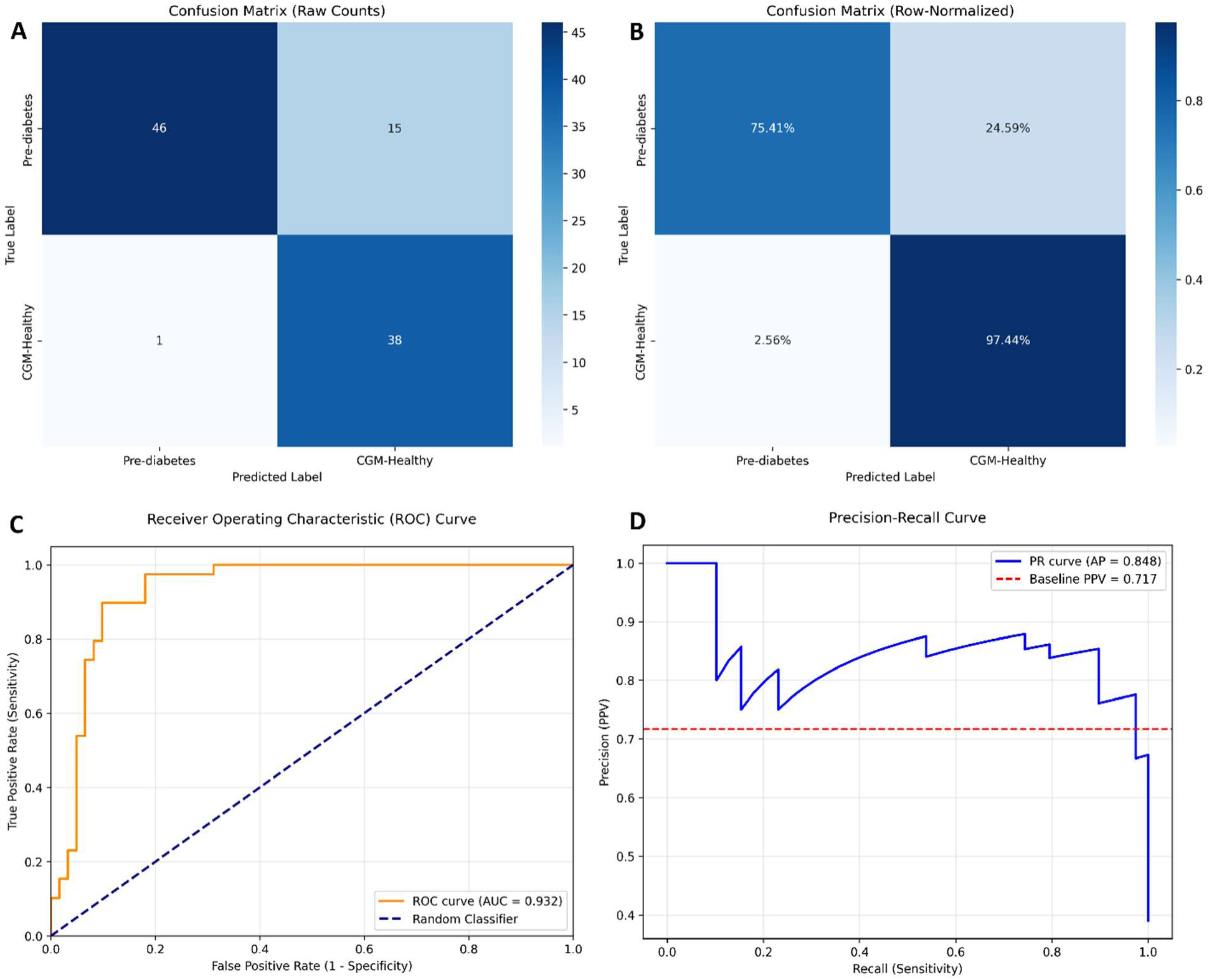
LSTM performance on the held-out Test set (predicted study groups: Healthy vs pre-diabetic). (a) Confusion matrix using counts. (b) Confusion matrix using row-normalized in percentages. (c) Receiver Operating Characteristic (ROC) curve, AUC of 0.88. (d) Precision (PPV) and Recall (sensitivity) across thresholds.

Calibration assessment confirmed that predicted probabilities accurately reflect true class membership likelihood like Brier Score showing 0.101, Expected Calibration Error (ECE) of 0.075 (<0.10 indicates excellent calibration), and temperature scaling T of 1.000, indicating the model was naturally well-calibrated and suitable for confidence-based clinical decision support.

The 3-tier confidence system, using dynamic thresholds centered on the global threshold (0.375), demonstrated practical clinical utility on the held-out test set. Zone 1 (high confidence prediabetes, probability < 0.295) contained 46 participants (46.0%) with precision ≈ 97.8% and false positive rate ≈ 2.6%, enabling immediate lifestyle intervention without confirmatory OGTT. Zone 2 (uncertain, probability 0.295–0.595) captured 6 participants (6.0%) requiring OGTT, representing the OGTT burden of 6.0%. Zone 3 (high confidence CGM-Healthy, probability ≥ 0.595) contained 48 participants (48.0%) with precision ≈ 77.1% and specificity ≈ 94.9%, allowing regular screening without immediate intervention. Overall, the system achieved detection rate ≈ 82.0% while maintaining low OGTT burden (6.0%) and low false positive rate (2.6%), providing actionable clinical recommendations with high confidence in most cases.

A key strength of this work is the potential to integrate the Conv+BiLSTM model (trained on rigorously cleaned labels) directly into CGM device interfaces, enabling real-time, continuous prediabetes risk assessment as users wear their sensors. This transforms passive glucose monitoring into an active early-warning system for metabolic health, providing immediate risk scores without requiring separate diagnostic visits. The Conv+BiLSTM classification model was integrated into the PredictMod platform^37^ as proof-of-concept for such deployment, demonstrating the feasibility of embedding advanced machine learning models within existing diabetes care workflows.

### Temporal Requirements Analysis

We note that 7 days of CDM sequence data were required for adequate training of the model. At 7 days, the model reached its peak performance, hitting a remarkable AUC of 0.93. After 7 days, the benefits started to level off, with only slight improvements noted. This analysis tells us that for accurate classification, the algorithm requires at least 7 days of CGM data, which represents a feasible balance between accuracy and patient inconvenience. This study supports a possible clinical deployment in which extensive monitoring (>2 weeks) may impact how well patients comply.

### Feature Importance and Clinical Insights

To understand how the Conv+BiLSTM model makes predictions and which glucose features most strongly discriminate between CGM-Healthy and prediabetes states, we performed sequence-aware permutation importance analysis stratified by predicted class (Fig. 5). This technique measures how much model performance (AUC) degrades when each feature is randomly shuffled (permuted) separately for samples predicted as Pre-Diabetic (red bars) versus samples predicted as CGM-Healthy (blue bars). The key insight is that positive bars indicate features that help identify that class (AUC drops when permuted) and negative bars indicate features supporting the opposite class. The opposite-direction pattern (red vs. blue bars) reflects the binary nature of classification.

**Figure 5.**
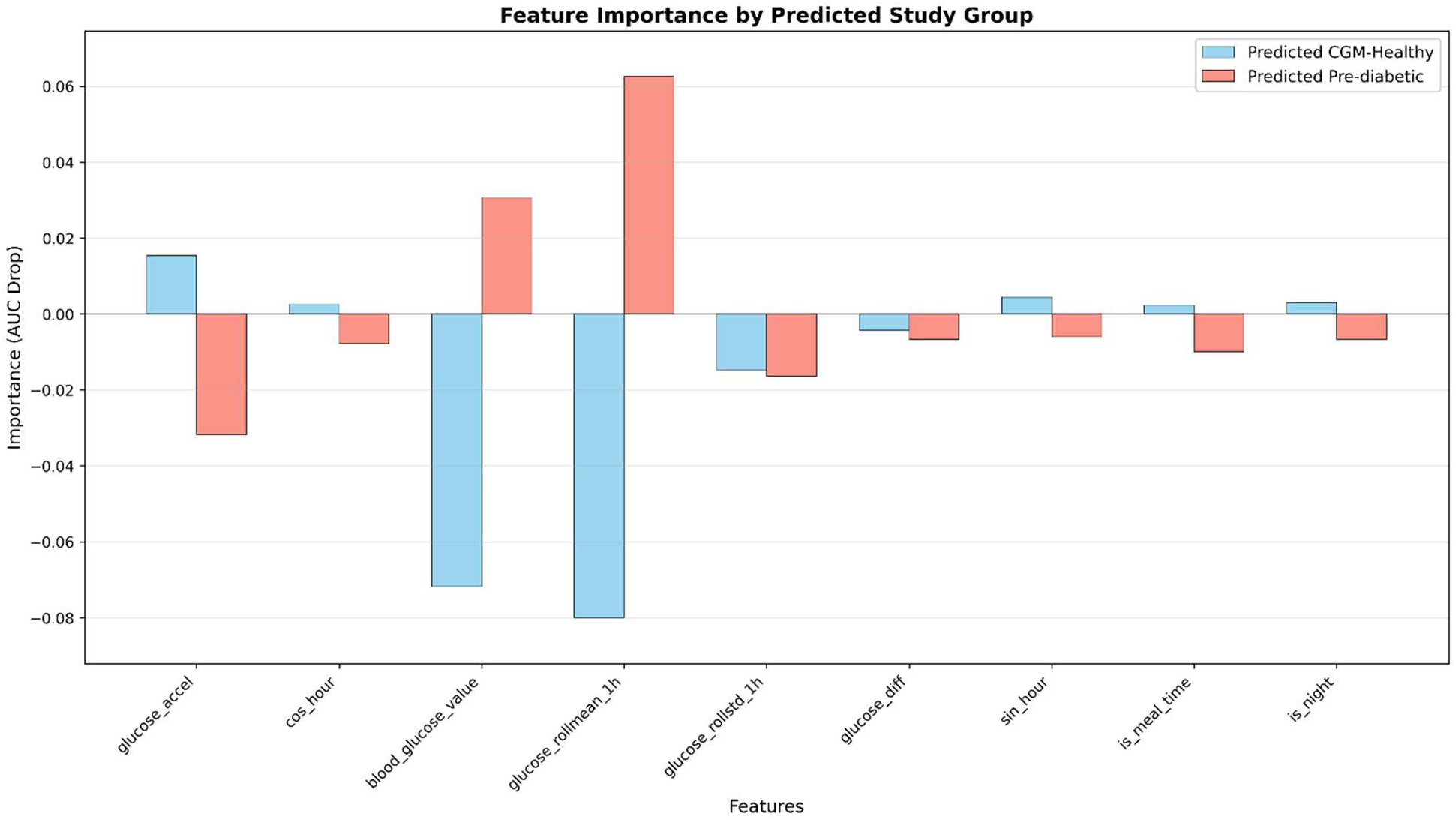
Feature importance by predicted study group (healthy vs. pre-diabetic). Sequence-aware permutation importance (ΔAUC) stratified by predicted study group; higher ΔAUC denotes greater contribution.

Figure 5 shows glucose_rollmean_1h feature emerged as the strongest discriminator, with the largest magnitude bars in opposite directions (Pre-Diabetic: +0.062, CGM-Healthy: −0.080), indicating that elevated short-term average glucose is the most powerful predictor of prediabetes while consistently low rolling mean glucose strongly indicates healthy glycemic control. This aligns with clinical understanding that sustained hyperglycemia, even within sub-diabetic ranges, is a hallmark of prediabetes, and the 1-hour window effectively captures postprandial glucose handling, which deteriorates early in diabetes progression. blood_glucose_value feature showed moderate importance with the model relying more on temporal averaging than instantaneous readings. glucose_accel feature revealed an interesting opposite-directional pattern, indicating CGM-Healthy individuals exhibit rapid corrective acceleration after glucose excursions while Pre-Diabetic individuals show sluggish, irregular acceleration, aligning with known β-cell dysfunction and impaired insulin response in early diabetes progression. Circadian and temporal features showed minimal class-specific effects. These findings confirm that the Conv+BiLSTM model has learned physiologically meaningful patterns rather than spurious correlations, increasing confidence in its predictions and potential for clinical deployment. The model essentially identifies prediabetes through sustained hyperglycemia, elevated glucose spikes, and impaired corrective responses, the core metabolic signatures of early diabetes.

Multicollinearity diagnostics revealed high correlation between glucose_rollmean_1h and blood_glucose_value (r = 0.996, Variance Inflation Factor (VIF) ≈ 7.7 for both features), indicating redundancy. However, both features were retained as they capture complementary temporal scales: blood_glucose_value represents instantaneous measurements, while glucose_rollmean_1h smooths over 1-hour windows to capture meal-response dynamics. All continuous features exhibited VIF < 10 (acceptable multicollinearity threshold), confirming model stability despite feature correlation. Healthy patients showed a quick return to normal glucose levels after meals, usually within two (<2) hours. They also had minimal overnight glucose changes and consistent daily glucose patterns. In comparison, prediabetes individuals went through extended glucose fluctuations after eating, lasting longer than three (>3) hours. Their nighttime glucose levels fluctuated more, and their daily rhythms were irregular.

### Analysis of Heart Rate, Activity Level, and Sleep Data

Additionally, we analyzed heart rate, activity level, and stress and sleep data from the AI-READI dataset and validated it against CGM data to evaluate the predictive value for glycemic status. Using both the original and optimized dataset labels and limiting to healthy and prediabetic patients, we trained ensemble bagged decision tree models. Validation accuracy improved significantly when using the cleaned labels (from 57.1% to 82.2%) and ROC analysis confirmed stronger discrimination (AUC 0.52 vs. 0.61). Confusion matrices indicated reduced misclassification with optimized labels. Model inference supported evidence that heart rate, activity, and stress and sleep measures were consistently influential. Partial dependence plots suggested that vigorous cardiovascular activity and low stress were strongly associated with healthy status. Local Shapley^67^ explanations further illustrated how individual-level combinations of exercise and stress shaped predictions. However, despite these promising findings, the models only achieved moderate accuracy, reflecting both the complexity of diabetes pathophysiology and the challenges of limited wearable data streams. Future studies should extend observation windows beyond daily summaries to capture longitudinal patterns and test generalizability across diverse cohorts. Our results indicate that wearable-derived data provide clinically interpretable signals of metabolic health and, when validated against CGM data, can serve as scalable tools for predictive screening and early intervention.

## DISCUSSION

### Principal Findings and Clinical Significance

The present study demonstrates a substantive change in what we learn from CGM with label-aware pre-processing. Among participants originally labeled as “healthy”, (n=283), 43.1% (122 participants) displayed glucose profiles consistent with normative physiology (high TIR, low variability), while 56.9% (161 participants) demonstrated glucose profiles consistent with prediabetes. After an unsupervised phenotype audit and an iterative, expert-in-the-loop relabeling step, we were able to derive a cleaned cohort to which the downstream Conv+BiLSTM demonstrated excellent discrimination showing a rise in ROC-AUC of 0.025 from 5-fold cross-validation to testing on held-out test set with sine/cosine circadian encodings and nine physiologically grounded time-series features including rolling statistics (mean, standard deviation), glucose derivatives (first and second derivatives), circadian encodings (sine/cosine hour), and temporal flags (meal time, night). Consolidating this audit → relabel → sequence-model pipeline reduced label discordance and enhanced face validity of the supervised task, implying that merging CGM-derived evidence with expert review is a feasible path to greater reliability of prediabetes classification. These results provide evidence in support of an important argument that curation, specifically reconciling self-report/HbA1c-derived labels with CGM-derived evidence, enables face validity and improved model performance for classification of early dysglycemia.

Initial LSTM architectures struggled with the long CGM sequences (2,138 timesteps, ≈7.4 days), exhibiting vanishing gradients and overfitting despite regularization attempts. The Conv+BiLSTM hybrid architecture addressed these limitations through a two-stage approach: convolutional layers first compress the sequence from 2,138 to ≈133 timesteps while extracting local temporal patterns (meal responses, glucose excursions), bringing the sequence within the effective gradient-propagation range of LSTMs; bidirectional LSTM layers then capture long-range dependencies by processing temporal context in both forward and backward directions. This architectural innovation enabled the model to learn from week-long glucose trajectories without information loss or gradient instability, a critical requirement for detecting subtle prediabetes patterns that manifest over multi-day timescales. The resulting model demonstrated robust generalization across cross-validation folds, with consistent performance (ROC-AUC standard deviation <0.03) indicating minimal overfitting, a stark improvement over earlier LSTM-only attempts that showed substantial fold-to-fold variability.

Beyond classification accuracy, there is a practical contribution to the literature of a recipe for operationalizing clinical priors in CGM data. The clustering step formalized what clinicians would expect to see in a CGM-H trace (high TIR, stable profiles overnight, brisk postprandial decay), whereas the sequence model captured dynamic elements (rates and accelerations, circadian structure, meal windows) that capture the variability in predictions across individuals. The performance noticeably saturated at 7 days of data, which also serves to appropriately balance clinical feasibility and overall signal sufficiency. This aligns with the idea that early dysglycemia is better recognized from patterns like how fast glucose rises and cools, how variable nights are, rather than from a single static marker, and it suggests a concrete monitoring horizon for screening use-cases.

### Cooling-Period Insights

Our analysis during the cooling period provides a mechanistic picture, formally using participant-specific “baselines” and personalized “living ranges” to define spikes, and to demand sustained returns (≥15 minutes) to assess recovery. In cleaned CGM-H profiles, recoveries clustered around clinically expected values while prolonged or censored recoveries were common in prediabetes, descriptively supporting our postprandial decay being slower. Shorter validation windows or permissive prominence settings can label shoulder peaks as distinct spikes and artificially inflate cooling times, and establishing detection from ranges and requiring sustained returns helps mitigate against these imposed artifacts. For future clinical translation, we recommend reporting for both (i) the median sustained return time, and (ii) the percent of runs where spikes were censored (no return in the 6-hr window) as complementary indicators of stability.

There are a number of strengths to this work. First, the model treats label quality as a first-class priority rather than as an afterthought, leveraging unsupervised discovery and structured review by experts. Second, it generates per-participant sequences that use masking rather than heavy imputation, which makes the modeling unit consistent with the mental model for patient-related thinking employed by clinicians. Third, the features and endpoints are intentionally interpretable (TIR, %CV, recovery time) and are usable for clinical auditability as well as prospective use. Finally, the model and pipeline are modular, each part of the pipeline (phenotype audit, relabel loop, per-participant sequence model, cooling analysis) can either be easily swapped out, tuned, or both, and it can be independently validated.

### Limitations

Several notable limitations exist. First, label adjudication relies on CGM-derived metrics; even though the LSTM leverages different inputs than clustering features, the risk of construct circularity remains (i.e., labels and features share a domain). Utilizing an external ground truth (e.g., standardized meal tests, oral glucose tolerance test) would alleviate some of that risk. Second, meal windows and sleep were defined heuristically; more accurate annotations of real-life events (e.g., meals, exercise, medications) will improve spike attribution and cooling endpoints for accurate definitions. Third, SMOTE was applied only in the tabular XGBoost relabeling loop (train-only), whereas the Conv+BiLSTM time-series model employed class-weighted training without oversampling. Fourth, we have not yet tested generalizability beyond device and dataset. We excluded traces with strings “LOW/HIGH” and shorter traces; these exclusions may result in bias towards clearer signals for glucose patterns. Fifth, the expert review step, which facilitated valuable input, introduces the risk of reviewer bias; multi-rater adjudication with inter-rater agreement (e.g., Cohen’s κ^68^) should be used to increase reliability. Sixth, while Conv+BiLSTM addressed overfitting challenges inherent in long-sequence modeling, the architecture’s complexity (121,729 parameters) may limit interpretability compared to simpler models. Future work should explore attention mechanisms or hybrid approaches that maintain performance while offering greater transparency into feature importance and decision pathways. Lastly, the study was focused on binary classification (CGM-H vs prediabetes), and extending this work to multiclass classification or ordinal stages will be important to clinical implementation.

### Future Directions

Future research should include prospective and external validation. Feasible next steps are to use a standardized meal challenge cohort with concurrent capillary measurements, to calibrate postprandial kinetics, to characterize the interstitial lag associated with CGM, and to verify cooling endpoints. In terms of methodology, employing probability-based audits on labels at some of the cross-validated confident-learning style error estimates, using the human-in-the-loop, sequence-aware augmentation process, and auditing the fairness of models across age, sex, and body mass index (BMI) strata would improve their robustness. Disease progression might be better characterized by moving beyond simple binary classification to a multiclass or ordinal scale (healthy → prediabetes → medication-treated states), especially when combined with future multimodal information streams (i.e., activity, ECG) from AI-READI. For clinical implementation, decision thresholds should be calibrated around use cases (screening vs. triage), and output of the work should be supplemented with patient-level results that summarize aspects of TIR (%CV, median cooling, censored fraction) to facilitate clinical decision-making.

## Conclusion

By standardizing sequence-aware, clinically interpretable features and establishing data requirements ≈7 days, we have outlined a practical route to deployable CGM analytics. A label-aware, expert-in-the-loop pipeline that reconciles survey/HbA1c labels with CGM evidence provides marked improvements in validity and prediabetes classification accuracy. The approach is modular and generalizable to other biosensor datasets, where label noise and the temporal sequence of data representation are primary considerations, and may further facilitate earlier identification of dysglycemia and the provision of targeted intervention.

## Data Availability

Data are publicly available via the Bridge2AI AI-READI project (https://aireadi.org/).

## Abbreviations

CDC: Centers for Disease Control
CGM: Continuous glucose monitoring
TIR: Time in Range
TAR: Time Above Range
TBR: Time Below Range
ATTD: Advanced Technologies & Treatments for Diabetes
ADA: American Diabetes Association
GV: Glycemic Variability
%CV: Coefficient of Variation of Glucose
MAGE: Mean Amplitude of Glycemic Excursions
CONGA: Continuous Overlapping Net Glycemic Action
CGM-H: CGM healthy
SD: Standard Deviation
LSTM: Long Short-Term Memory
AI-READI: Artificial Intelligence Ready and Exploratory Atlas for Diabetes Insights
NIH: National Institutes of Health
HbA1c: Hemoglobin A1
PCA: Principal Component Analysis
SMOTE: Synthetic Neighbor Oversampling Technique
AUC: Area Under the Curve
ROC: Receiver Operating Characteristic
MCC: Matthews Correlation Coefficient
ECE: Expected Calibration Error
IQR: Interquartile Range
BA: Balanced Accuracy
BMI: Body Mass Index
VIF: Variance Inflation Factor
OGTT: Oral Glucose Tolerance Test

## ACKNOWLEDGEMENTS

We would like to acknowledge the numerous graduate students, interns, and volunteers for their dedicated efforts.

